# Blood Pressure Control From 2011 To 2019 In Patients 90 Days After Stroke

**DOI:** 10.1101/2023.02.12.23285827

**Authors:** Deborah A. Levine, Lewis B. Morgenstern, Madeline Kwicklis, Xu Shi, Erin Case, Lynda D. Lisabeth

## Abstract

**Background and Aims:** Whether and how much stroke survivors’ blood pressure (BP) control changed over the 2010s decade is unclear. We assessed whether 90-day BP control changed from 2011 to 2019, and whether temporal changes varied by ethnicity and sex.

**Methods:** We conducted a prospective cohort study of 1,755 first expert-adjudicated stroke cases ≥45 years in the Brain Attack Surveillance in Corpus Christi (BASIC) project with BP measurements from 2011-2019. We measured BP in patients’ residences at ∼90 days post-stroke using standardized procedures and validated oscillometric devices. The average of three BP measurements was used. Outcomes were BP control <130/80 mmHg (primary) and BP control<140/90 mmHg (secondary). We used inverse probability weights to account for attrition due to 90-day mortality and non-participation in study interviews. Using logistic regression, we examined the association between BP control and time (year) and demographic differences in trends through interactions with time adjusting for patient factors.

**Results:** Median age was 66 years (interquartile range, 58-76 years), 59% were Mexican American, 49% were women. From 2011 to 2019, BP control <130/80 mmHg declined from 43.0% to 28.6% (P<0.001). Odds of BP control <130/80 mmHg decreased over time (odds ratio per one-year increase, 0.95; 95% CI, 0.91, 0.99) after adjustment for patient factors. BP control <140/90 mm Hg remained consistent at 59.3% in 2011 and 57.1% in 2019 (P=0.31). Odds of BP control <140/90 mm Hg appeared constant over time after adjustment for patient factors (odds ratio per one-year increase, 1.00; 95% CI, 0.96, 1.04). Trends in BP control <130/80 mmHg and <140/90 mmHg did not significantly differ by ethnicity or sex.

**Conclusions:** From 2011 to 2019, BP control <130/80 mmHg decreased and BP control <140/90 mmHg did not improve in patients 90 days after stroke. Results suggest stroke survivors need effective, sustainable strategies to achieve BP control.

**AHA Journals Subject Terms:** Cerebrovascular Disease/Stroke, Blood Pressure, Quality and Outcomes, Health Services

## Introduction

Stroke survivors have increased risk for mortality and cardiovascular events, with the risk of recurrent stroke highest during the first 90 days after stroke. Blood pressure (BP) control reduces these adverse outcomes. Knowing the temporal trends in BP control 90 days after stroke is critical to understanding the quality of stroke survivors’ care and informing interventions to improve care. It is unknown how BP control 90 days after stroke changed from 2011 to 2019. Prior studies of BP control in US stroke survivors have lacked data after 2016.^1-3^ The proportion of US stroke survivors with BP control could have decreased after 2016 as uninsurance and cost-sharing have increased.^4, 5^

Temporal changes in BP control after stroke might vary by sex and Hispanic ethnicity. Studies suggest that women and Hispanic individuals are more likely to have uncontrolled BP after stroke.^2^ Yet, it is unknown whether temporal changes in BP control 90 days after stroke during the 2010s vary by sex and Hispanic ethnicity. Identifying stroke survivor subgroups who have worsening BP control over time will provide data to support targeted interventions for those who need them most.

Leveraging a well-characterized, US cohort of non-Hispanic White and Mexican American stroke cases, we assessed whether 90-day BP control after stroke changed from 2011 to 2019, and whether temporal changes varied by ethnicity and sex. We hypothesized that 90-day BP control after stroke decreased from 2011 to 2019, with greater decreases in women than men and Hispanic individuals than non-Hispanic White individuals.

## Methods

Study methods are reported adhering to STROBE guidelines for observational studies.

### Study Population and Recruitment

The Brain Attack Surveillance in Corpus Christi (BASIC) project is a population-based stroke surveillance study conducted in a non-immigrant community of primarily Mexican Americans and non-Hispanic whites in Nueces County, Texas. Details are described elsewhere.^6^ Briefly, Nueces County is a predominantly urban location, where 95% of the population resides in the city of Corpus Christi on the Texas gulf coast. Corpus Christi is situated >150 miles from potential referral centers in San Antonio and Houston. The geographic location and distance provide the opportunity for complete case capture of stroke in the county.

Study coordinators identified stroke cases through active and passive surveillance using rigorous criteria. Stroke physicians validated stroke cases using source documentation following international clinical criteria. At the time of stroke hospitalization or soon after, patients (or proxies for patients unable to participate) completed an in-person, structured interview. Bilingual study coordinators conducted the interview in English (94%) or Spanish (6%) per patient preferences. Most of the Mexican American participants are second- and third-generation Americans. Interview participation was similar by ethnicity. This project was approved by both the University of Michigan and Corpus Christi Health Systems’ Institutional Review Boards. All subjects or their proxies provided informed consent.

We required participants to have measurements of BP at the 90-day in-person outcome assessment. We excluded participants who were unable to complete the baseline interview and 90-day outcome assessment.

We identified 1,914 BASIC participants with their first BASIC stroke (ischemic or hemorrhagic) in the study between March 2011 through December 2019 who completed the baseline interview and had complete outcome information. Of these, we excluded 159 individuals missing BP measurements due to phone interviews and refusal to do the BP measurement portion of the interview (Figure I in the Data Supplement). This project was approved by both the University of Michigan and Corpus Christi Health Systems’ Institutional Review Boards. All subjects or their proxies provided informed consent.

### Outcome Measurement

Study coordinators measured BP at the 90-day outcome assessment using a standard protocol and validated oscillometric devices.^7^ Following a 5-minute rest, BP was measured three times in the right arm of seated participants at 15-second intervals using an appropriately sized cuff and a standard automated BP measurement arm monitor (OmROn model 700 series; Omron, Mannheim, Germany), that has been validated by the Association for the Advancement of Medical Instrumentation to be accurate. The BP of record was the average of the 3 BP measurements. The primary outcome was BP control <130/80 mmHg because guidelines recommend this goal in stroke survivors^8^ and define hypertension as BP ≥130/80 mm Hg.^9^ The secondary outcome was BP control <140/90 mmHg.

### Covariates

Covariates were selected using the Andersen Behavioral Model for vulnerable populations and measured at baseline (time of index stroke) except where noted. Participants self-reported age, sex, race/ethnicity (non-Hispanic White, Mexican American, Other), education (<high school, high school, >high school), and marital status. National Institutes of Health Stroke Scale (NIHSS) score and co-morbidities were abstracted from the medical record. Comorbidities were hypertension, history of stroke/TIA, diabetes, coronary artery disease/myocardial infarction, atrial fibrillation, high cholesterol, excessive alcohol use, cigarette smoking, congestive heart failure (CHF), depression, and end-stage renal disease. Body mass index was calculated from height and weight obtained from the medical record. Stroke type (ischemic vs hemorrhagic) was classified by study neurologists.

The short form of the Informant Questionnaire on Cognitive Decline in the Elderly (IQCODE), a validated instrument to assess pre-stroke cognitive status^10-12^ was assessed by an informant. We used the IQCODE and medical record history of dementia/Alzheimer’s disease to categorize patients as normal cognition (IQCODE score ≤3), mild cognitive impairment (IQCODE score 3.1-3.43), and dementia (IQCODE score ≥3.44 or dementia/AD history).^7^ Access to care variables were self-report of a regular primary care physician and insurance status. Participants self-reported the number of limitations in basic and instrumental activities of daily living (ADL/IADL) at 90 days.

### Statistical Analysis

We followed a pre-specified analysis plan. Inverse probability weights were constructed using logistic regression to account for sample bias due to non-participation in baseline and outcome interviews and mortality before 90 days.^13^ Attrition from the three different sources was modeled separately;^14^ mortality was modeled conditioning on baseline participation, and outcome participation was modeled conditioning on baseline participation and survival to 90 days. Each set of resulting weights from the three models, computed as the reciprocal of the estimated probability of participation/survival, was stabilized. The three sets of weights were then multiplied together to determine the final set of weights.

The main models were estimated using logistic regression. The overall trend was quantified as the linear relationship between time and log odds. The Hosmer-Lemeshow goodness-of-fit test showed good model fit. Visual inspection of the predicted values vs. observed values showed good model calibration. A quadratic time term was not significant. We examined whether temporal trends in BP differed by race/ethnicity and sex through interactions with time (time*race/ethnicity, time*sex, and time*race/ethnicity*sex). Sex, age, education, marital status, comorbidities including history of hypertension, pre-stroke cognition, BMI, initial NIHSS, smoking status, regular primary care physician, insurance, ADL/IADL limitations at 90 days, stroke type (ischemic vs hemorrhagic), and history of stroke/TIA, were included as covariates. Missing covariates were imputed using the R package ‘mice’,^15^ and the 25 datasets generated by Multiple Imputation with Chained Equations (MICE) were pooled using Rubin’s rules.^16^ Analyses were performed using R v4.1.3.

## Results

Figure 1 shows the derivation of the cohort. Median age was 66 years (interquartile range, 58-76 years), 59% were Mexican American, 49% were women (Table 1).

**Figure 1:**
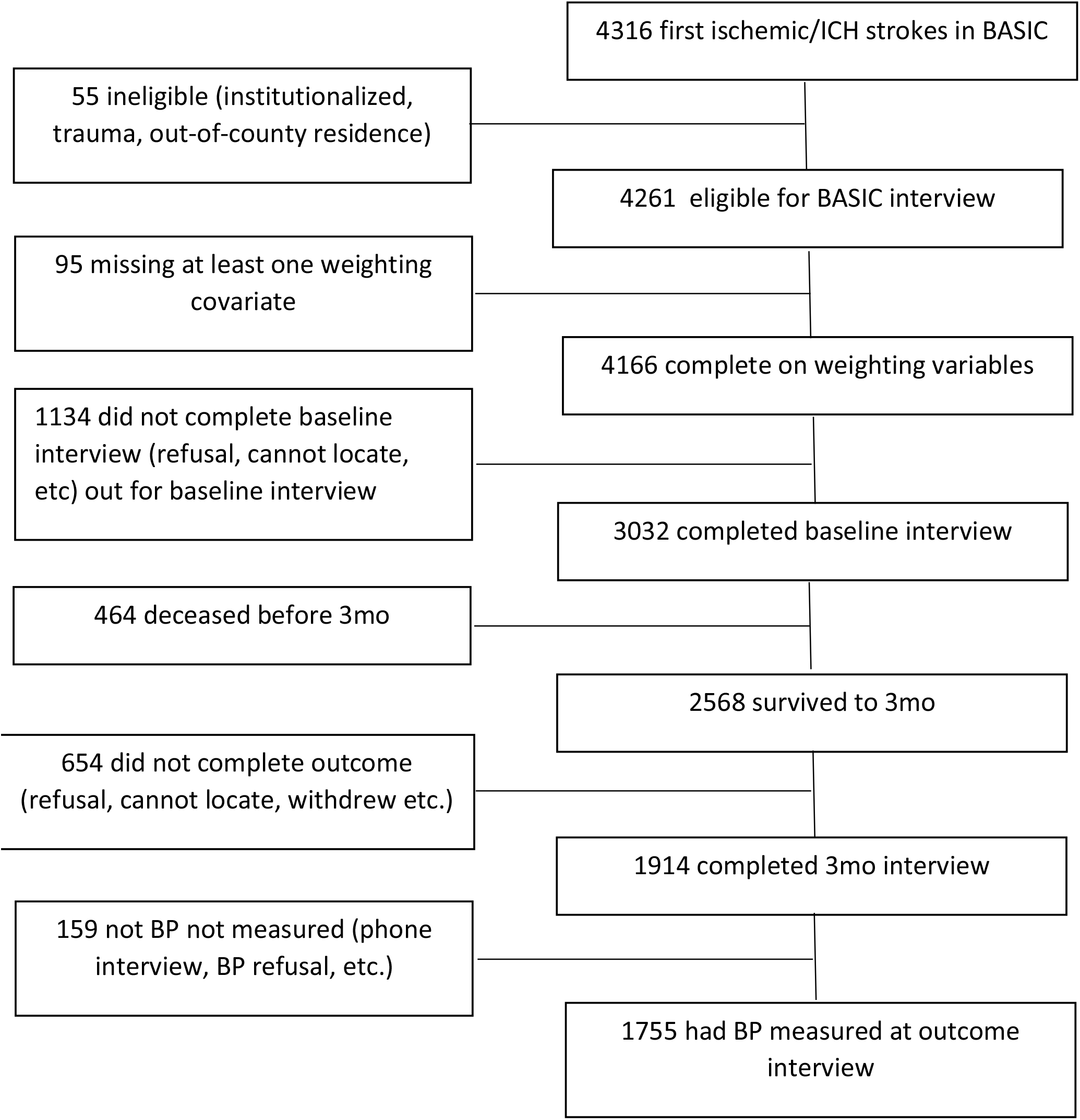
Derivation of the Study Sample: BASIC Project, 2011-2019.

**Table 1:**
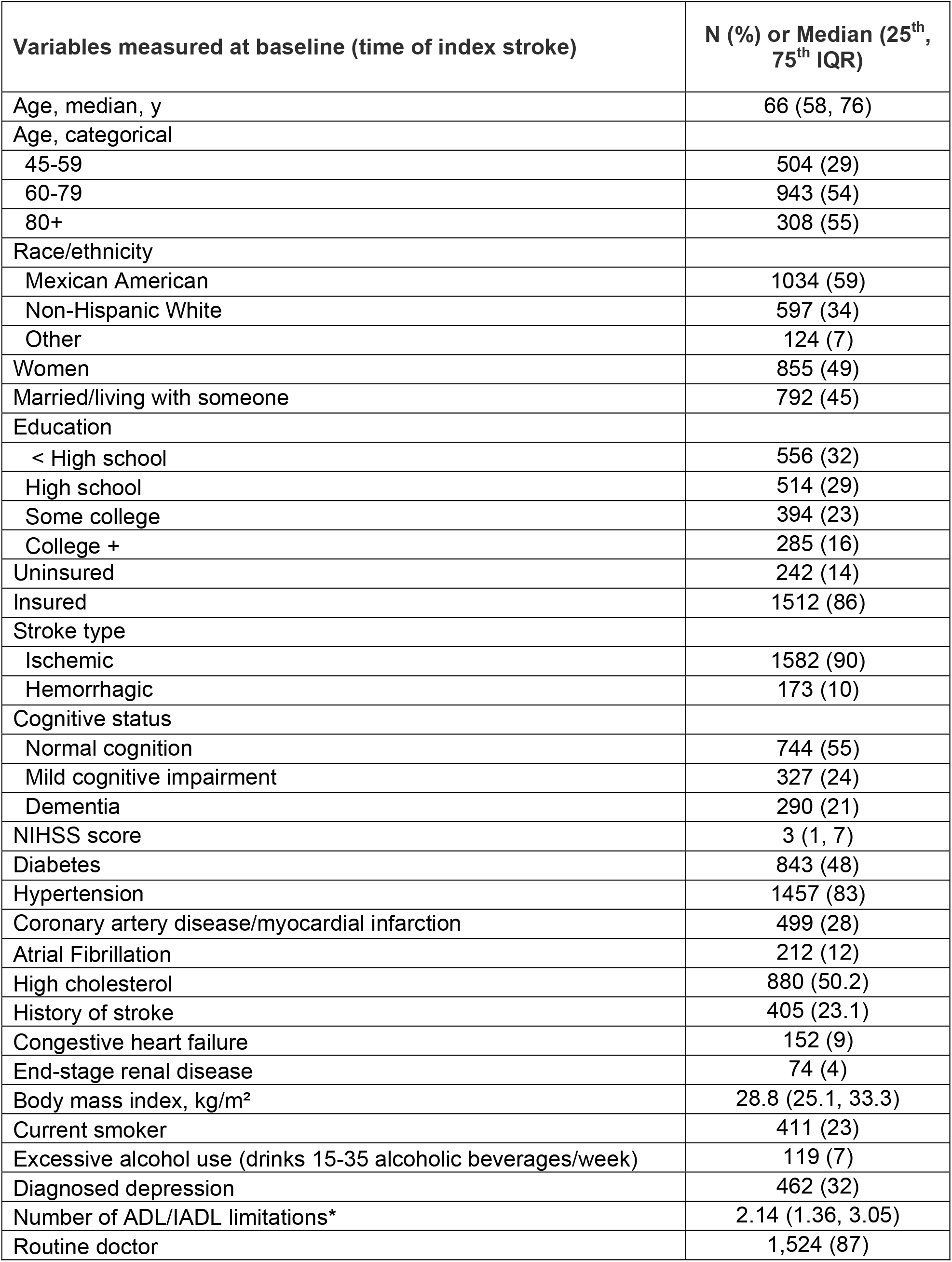
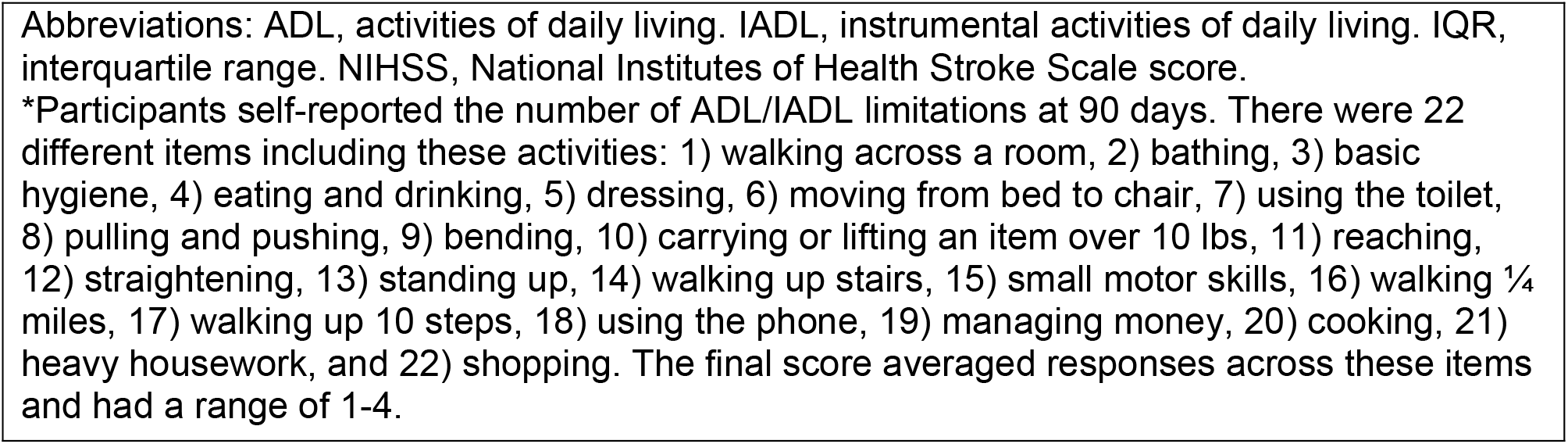
Characteristics in the Study Sample (n=1,755)

| **Variables measured at baseline (time of index stroke)** | **N (%) or Median (25<sup>th</sup>, 75<sup>th</sup> IQR)** |
| --- | --- |
| Age, median, y | 66 (58, 76) |
| Age, categorical |  |
| 45-59 | 504 (29) |
| 60-79 | 943 (54) |
| 80+ | 308 (55) |
| Race/ethnicity |  |
| Mexican American | 1034 (59) |
| Non-Hispanic White | 597 (34) |
| Other | 124 (7) |
| Women | 855 (49) |
| Married/living with someone | 792 (45) |
| Education |  |
| < High school | 556 (32) |
| High school | 514 (29) |
| Some college | 394 (23) |
| College + | 285 (16) |
| Uninsured | 242 (14) |
| Insured | 1512 (86) |
| Stroke type |  |
| Ischemic | 1582 (90) |
| Hemorrhagic | 173 (10) |
| Cognitive status |  |
| Normal cognition | 744 (55) |
| Mild cognitive impairment | 327 (24) |
| Dementia | 290 (21) |
| NIHSS score | 3 (1, 7) |
| Diabetes | 843 (48) |
| Hypertension | 1457 (83) |
| Coronary artery disease/myocardial infarction | 499 (28) |
| Atrial Fibrillation | 212 (12) |
| High cholesterol | 880 (50.2) |
| History of stroke | 405 (23.1) |
| Congestive heart failure | 152 (9) |
| End-stage renal disease | 74 (4) |
| Body mass index, kg/m<sup>2</sup> | 28.8 (25.1, 33.3) |
| Current smoker | 411 (23) |
| Excessive alcohol use (drinks 15-35 alcoholic beverages/week) | 119 (7) |
| Diagnosed depression | 462 (32) |
| Number of ADL/IADL limitations* | 2.14 (1.36, 3.05) |
| Routine doctor | 1,524 (87) |

Abbreviations: ADL, activities of daily living. IADL, instrumental activities of daily living. IQR, interquartile range. NIHSS, National Institutes of Health Stroke Scale score.
\*Participants self-reported the number of ADL/IADL limitations at 90 days. There were 22 different items including these activities: 1) walking across a room, 2) bathing, 3) basic hygiene, 4) eating and drinking, 5) dressing, 6) moving from bed to chair, 7) using the toilet, 8) pulling and pushing, 9) bending, 10) carrying or lifting an item over 10 lbs, 11) reaching, 12) straightening, 13) standing up, 14) walking up stairs, 15) small motor skills, 16) walking $\frac{1}{4}$ miles, 17) walking up 10 steps, 18) using the phone, 19) managing money, 20) cooking, 21) heavy housework, and 22) shopping. The final score averaged responses across these items and had a range of 1-4.

### Primary Outcome

From 2011 through 2019, BP control <130/80 mm Hg declined from 43.0% to 28.6% (P for trend <0.001) (Figure 2). Odds of BP control <130/80 mm Hg decreased over time after adjustment for socio-demographics, health insurance, regular primary care, clinical factors, and stroke features (odds ratio per one-year increase, 0.95; 95% CI, 0.91, 0.99) (Table 2). Trends in BP control <130/80 mm Hg did not differ by ethnicity (P=0.10) or sex (P=0.98).

**Figure 2:**
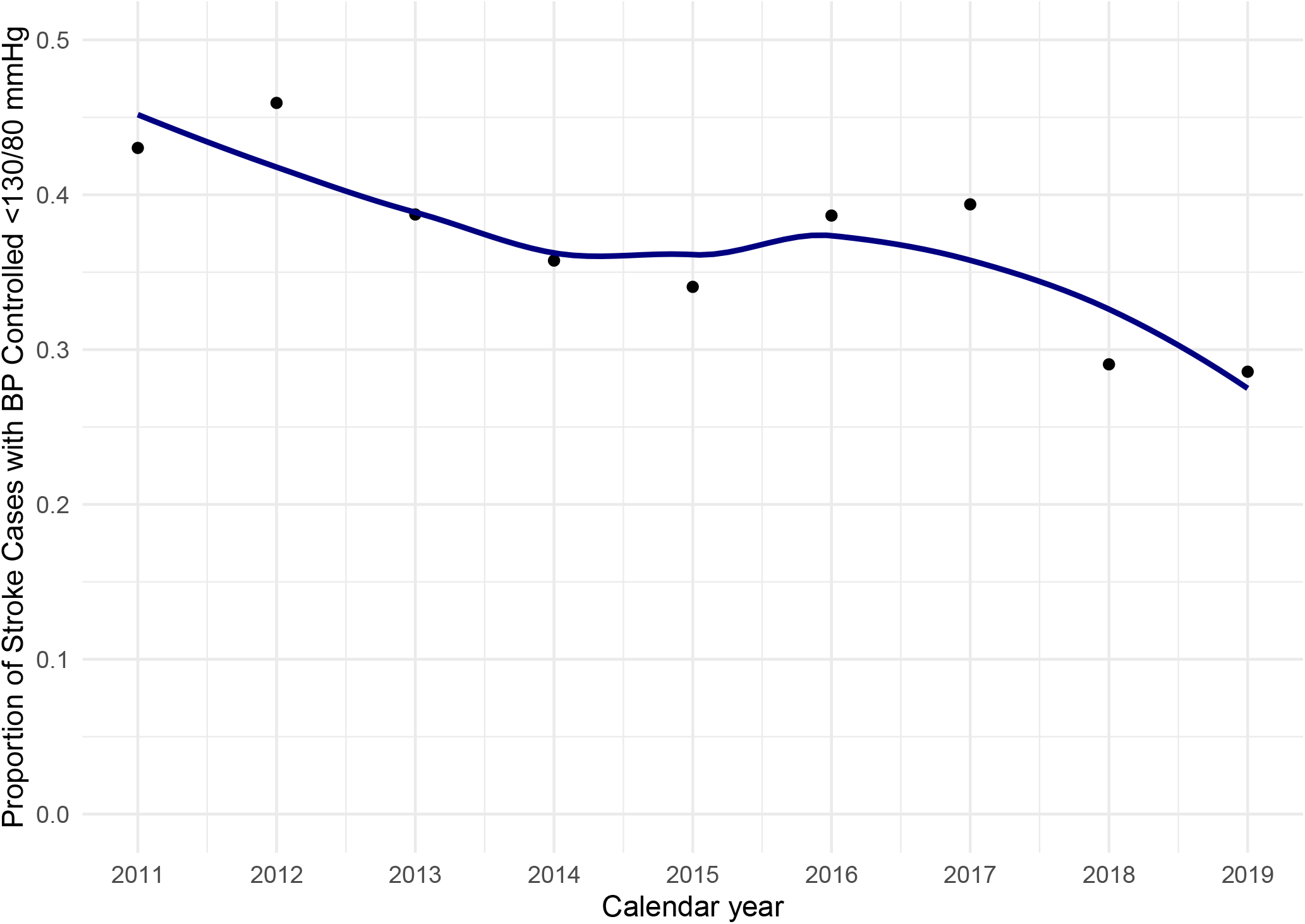
Proportion of Stroke Cases with BP Control <130/80 mmHg (Primary Outcome) 90 Days after Stroke over Time: BASIC Project, 2011-2019.

**Table 2:** Association between Patient Characteristics and Blood Pressure Control <130/80 mm Hg (Primary Outcome) 90 Days After Stroke.

|  | Adjusted odds ratio | 95% Confidence Interval | P-value |
| --- | --- | --- | --- |
| **Time per one-year increase** | **0.95** | **(0.91, 0.99)** | **0.01** |
| **Non-Hispanic White vs. Mexican American** | **1.35** | **(1.05, 1.74)** | **0.02** |
| Other race/ethnicity vs. Mexican American | 0.83 | (0.54, 1.27) | 0.39 |
| Female vs. male | 1.11 | (0.88, 1.39) | 0.38 |
| **Age 60-79 vs. 45-59** | **1.48** | **(1.13, 1.92)** | **<0.01** |
| **Age 80+ vs. 45-59** | **1.58** | **(1.09, 2.31)** | **0.02** |
| Married vs. unmarried | 1.05 | (0.85, 1.30) | 0.67 |
| Education |  |  |  |
| < High school vs high school | 0.95 | (0.71, 1.27) | 0.73 |
| Some college vs high school | 0.97 | (0.73, 1.29) | 0.83 |
| College+ vs high school | 0.85 | (0.61, 1.19) | 0.33 |
| **log(NIHSS+1) per one-unit increase** | **1.15** | **(1.03, 1.30)** | **0.02** |
| Hemorrhagic vs. ischemic stroke | 1.00 | (0.72, 1.37) | 0.98 |
| Diabetes | 1.00 | (0.79, 1.26) | 0.99 |
| Diagnosed hypertension | 0.80 | (0.60, 1.08) | 0.14 |
| Coronary artery disease/myocardial infarction | 1.05 | (0.82, 1.33) | 0.72 |
| Atrial fibrillation | 1.14 | (0.83, 1.57) | 0.40 |
| High cholesterol | 1.22 | (0.98, 1.53) | 0.08 |
| History of stroke | 0.85 | (0.66, 1.10) | 0.22 |
| **Congestive heart failure** | **1.58** | **(1.10, 2.27)** | **0.01** |
| End-stage renal disease | 0.88 | (0.53, 1.44) | 0.60 |
| BMI per one-unit increase | 0.98 | (0.97, 1.00) | 0.06 |
| Mild cognitive impairment vs. normal cognition | 1.10 | (0.83, 1.46) | 0.51 |
| Dementia vs. normal cognition | 0.98 | (0.72, 1.33) | 0.90 |
| Current smoker | 1.13 | (0.86, 1.49) | 0.37 |
| Drinks 15-35 alcoholic beverages/week vs. does not | 1.20 | (0.78, 1.86) | 0.41 |
| Diagnosed depression | 0.99 | (0.74, 1.31) | 0.94 |
| **No. of ADL/IADL limitations per one unit increase** | **1.25** | **(1.09, 1.42)** | **<0.001** |
| Routine doctor vs. no routine doctor | 1.06 | (0.75, 1.51) | 0.72 |
| Insured vs. uninsured | 1.20 | (0.84, 1.70) | 0.31 |
| Abbreviations: ADL, activities of daily living. IADL, instrumental activities of daily living.<br>NIHSS, National Institutes of Health Stroke Scale score.<br>**Bolded figures indicate 95% confidence interval does not include 1.0.** |  |  |  |

Stroke survivors who had age 45-59 and Mexican American ethnicity were more likely to have uncontrolled BP ≥130/80 mm Hg (Table 2). Whereas stroke survivors who had age 60+, non-Hispanic White ethnicity, higher initial NIHSS, CHF, and higher ADL/IADL limitations were more likely to have controlled BP <130/80 mm Hg.

### Secondary Outcome

BP control <140/90 mm Hg remained consistent at 59.3% in 2011 and 57.1% in 2019 (P for trend=0.31) (Figure 3). 60.4% of patients with hypertension according to the first definition (>130/80 mm Hg) also had BP >140/90 mm Hg. Odds of BP control <140/90 mm Hg appeared constant over time after adjustment for socio-demographics, health insurance, regular primary care, clinical factors, and stroke features (odds ratio per one-year increase, 1.00; 95% CI, 0.96, 1.04) (Table 3). Trends in BP control <140/90 mm Hg did not differ by ethnicity (P=0.82) or sex (P=0.87).

**Figure 3:**
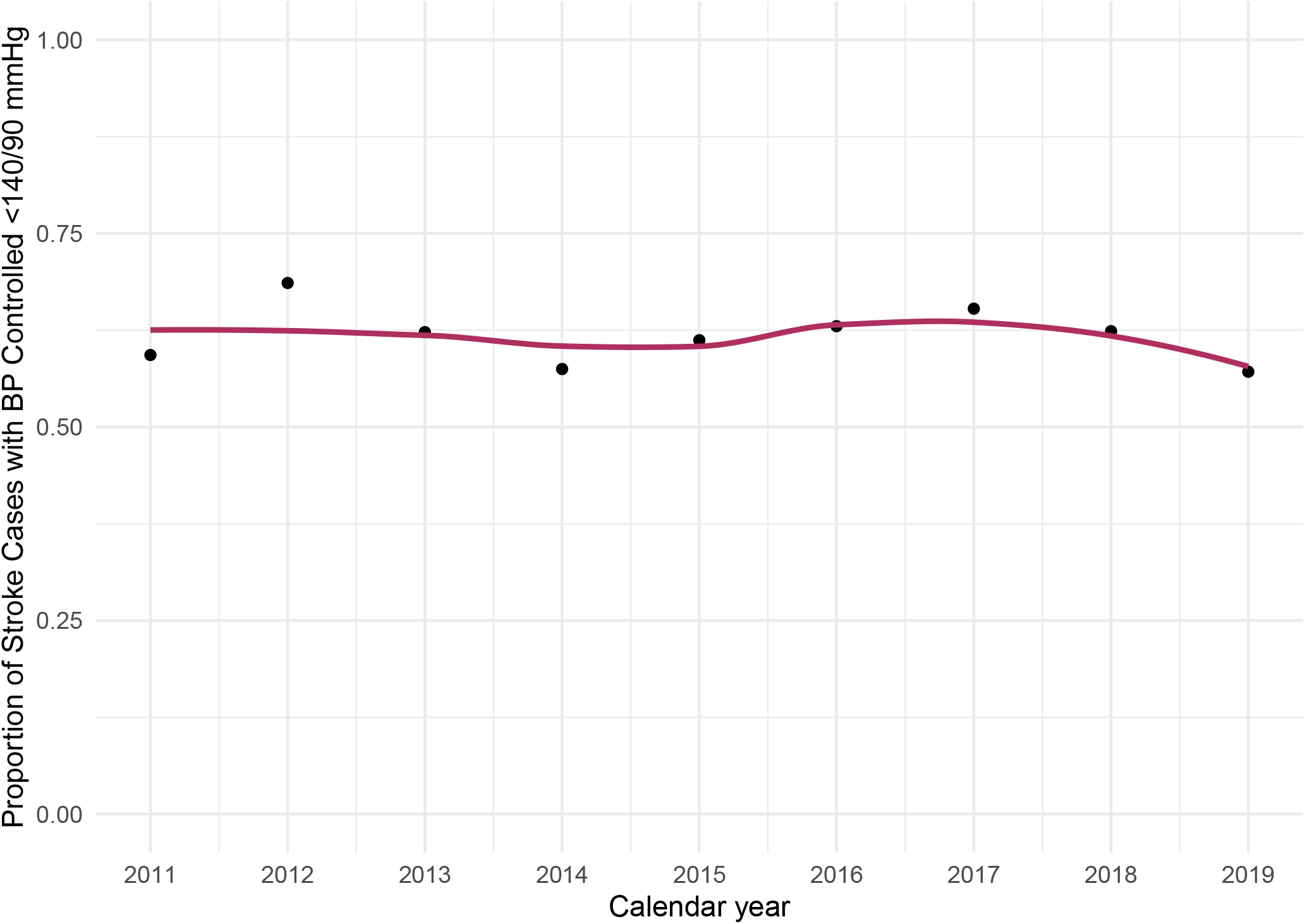
Proportion of Stroke Cases with BP Control <140/90 mmHg (Secondary Outcome) 90 Days after Stroke over Time: BASIC Project, 2011-2019.

**Table 3:** Association between Patient Characteristics and Blood Pressure Control <140/90 mm Hg (Secondary Outcome) 90 Days After Stroke.

|  | Adjusted odds ratio | 95% Confidence Interval | P-value |
| --- | --- | --- | --- |
| **Time per one-year increase** | 1.00 | (0.96, 1.04) | 0.90 |
| Non-Hispanic White vs. Mexican American | 1.27 | (0.98, 1.64) | 0.069 |
| **Other race/ethnicity vs. Mexican American** | **0.65** | **(0.44, 0.96)** | **0.033** |
| Female vs. male | 1.04 | (0.83, 1.3) | 0.72 |
| Age 60-79 vs. 45-59 | 1.15 | (0.89, 1.48) | 0.28 |
| Age 80+ vs. 45-59 | 0.91 | (0.62, 1.32) | 0.61 |
| **Married vs. unmarried** | **1.34** | **(1.08, 1.66)** | **< 0.01** |
| Education |  |  |  |
| < High school vs high school | 1.05 | (0.79, 1.38) | 0.75 |
| Some college vs high school | 1.17 | (0.87, 1.55) | 0.30 |
| College+ vs high school | 0.81 | (0.59, 1.13) | 0.22 |
| **log(NIHSS+1) per one-unit increase** | **1.18** | **(1.05, 1.33)** | **< 0.01** |
| Hemorrhagic vs. ischemic stroke | 1.04 | (0.75, 1.44) | 0.81 |
| Diabetes | 0.85 | (0.68, 1.07) | 0.17 |
| **Diagnosed hypertension** | **0.64** | **(0.48, 0.87)** | **< 0.01** |
| Coronary artery disease/myocardial infarction | 0.98 | (0.76, 1.25) | 0.86 |
| **Atrial fibrillation** | **1.43** | **(1.01, 2.01)** | **0.042** |
| High cholesterol | 1.02 | (0.82, 1.27) | 0.87 |
| History of stroke | 0.96 | (0.75, 1.24) | 0.77 |
| Congestive heart failure | 1.31 | (0.89, 1.92) | 0.18 |
| **End-stage renal disease** | **0.60** | **(0.37, 0.99)** | **0.045** |
| BMI per one-unit increase | 0.99 | (0.97, 1.01) | 0.22 |
| Mild cognitive impairment vs. normal cognition | 1.12 | (0.84, 1.48) | 0.44 |
| Dementia vs. normal cognition | 0.85 | (0.64, 1.14) | 0.29 |
| Current smoker | 1.25 | (0.96, 1.62) | 0.095 |
| Drinks 15-35 alcoholic beverages/week vs. does not | 0.97 | (0.64, 1.48) | 0.90 |
| Diagnosed depression | 1.05 | (0.81, 1.37) | 0.71 |
| **No. of ADL/IADL limitations per one unit increase** | **1.18** | **(1.03, 1.35)** | **0.016** |
| Routine doctor vs. no routine doctor | 1.34 | (0.96, 1.86) | 0.083 |
| Insured vs. uninsured | 1.31 | (0.95, 1.8) | 0.099 |
| Abbreviations: ADL, activities of daily living. IADL, instrumental activities of daily living.<br>NIHSS, National Institutes of Health Stroke Scale score.<br>**Bolded figures indicate 95% confidence interval does not include 1.0.** |  |  |  |

Stroke survivors who had other Mexican American ethnicity, diagnosed hypertension, and end-stage renal disease were more likely to have uncontrolled BP ≥140/90 mm Hg (Table 3). Conversely, stroke survivors who were married and had higher initial NIHSS, atrial fibrillation, and higher ADL/IADL limitations were more likely to have controlled BP <140/90 mm Hg.

## Discussion

In a population-based study of stroke survivors, we found that BP control <130/80 mm Hg assessed 90 days after stroke decreased substantially from 43.0% in 2011 to 28.6% in 2019. BP control <140/90 mm Hg 90 days post-stroke remained consistent at 59.3% in 2011 and 57.1% in 2019. We found no evidence that temporal trends in BP control using either definition differed by ethnicity or sex.

Prior studies were restricted to stroke survivors with a diagnosis of hypertension at or before 2016. Among individuals with stroke and hypertension, the frequency of both antihypertensive medication use and BP control <140/90 mm Hg remained constant from 2005 through 2016.^3^ However, many patients have undiagnosed hypertension at the time of stroke. Our study provides new evidence on the quality of BP control in unselected stroke survivors with and without a diagnosis of hypertension and including data after 2016. We found that odds of BP control <130/80 mm Hg decreased and odds of BP control <140/90 mm Hg did not improve from 2011 to 2019 after adjustment for patient factors including socioeconomic status, health insurance, regular primary care, clinical factors, and stroke features.

While BP control <130/80 mm Hg worsened from 2011 to 2019, stroke recurrence rates declined in the BASIC study from 2000 to 2013, continuing a previous trend.^17^ We hypothesize potential explanations for this paradox. The small worsening in BP control <130/80 mm Hg between 2011 and 2013 in BASIC (Figure 2) might not have had a large or immediate impact on stroke recurrence rates. The stability in BP control <140/90 mm Hg between 2011 and 2013 in BASIC (Figure 3) supports this theory. The persistent decline in stroke recurrence rates after adjusting for hypertension prevalence also suggests that non-BP factors might drive the trend.^17^ Stroke recurrence rates might have even been lower if BP was better controlled.

Our study has clinical and policy implications. Health systems should implement effective, multi-faceted, sustainable interventions to improve stroke survivors’ BP control.^18, 19^ Self-measured BP monitoring (SBPM) at home increases BP control and is cost-effective.^20^ Clinical guidelines recommend SBPM for stroke survivors to optimize BP control.^20^ However, many payers, including Medicare, do not cover SBPM devices except in patients on dialysis.^20^ To improve stroke survivors’ BP control, payers should provide SBPM devices, clinicians should recommend SBPM and provide feedback, and patients should share home BP readings with clinicians. Home SBPM ownership, clinician recommendation, and sharing BP readings with clinicians are associated with patients’ regular SBPM.^21^

Our study has limitations. We adjusted for education, insurance status, and primary care physician, but did not have information on other socioeconomic factors (e.g., income, wealth). We did not adjust for anti-hypertensive medication adherence because we did not have information on medication adherence for those without diagnosed and treated hypertension and our goal was to quantify BP control in all stroke survivors. Stroke survivors who had older age, non-Hispanic White ethnicity, and more severe strokes were more likely to be excluded; however, this would reduce our ability to find temporal changes in controlled BP because these survivors were more likely to have controlled BP and we used inverse probability weighting to minimize selection issues.

## Conclusion

From 2011 to 2019, BP control <130/80 mm Hg in patients 90 days after stroke decreased substantially and BP control <140/90 mm Hg did not improve. Results suggest stroke survivors require effective, sustainable solutions to control BP.

## Data Availability

Data referred to in this manuscript is the property of The Brain Attack Surveillance in Corpus Christi (BASIC) project. Data will be made available by request to the corresponding author pending approval of the BASIC study team.

## Acknowledgements

This study was performed in the Corpus Christi Medical Center and CHRISTUS Spohn Hospitals, CHRISTUS Health System, in Corpus Christi, Texas. The content is solely the responsibility of the authors and does not necessarily represent the official views of the National Institutes of Health (NIH).

## References

1. Sonawane K, Zhu Y, Balkrishnan R, Suk R, Sharrief A, Deshmukh AA, Aguilar D. Antihypertensive drug use and blood pressure control among stroke survivors in the United States: NHANES 2003-2014. J Clin Hypertens (Greenwich). 2019;21:766–773

2. Razmara A, Ovbiagele B, Markovic D, Towfighi A. Patterns and predictors of blood pressure treatment, control, and outcomes among stroke survivors in the United States. J Stroke Cerebrovasc Dis. 2016;25:857–865

3. Santos D, Dhamoon MS. Trends in antihypertensive medication use among individuals with a history of stroke and hypertension, 2005 to 2016. JAMA Neurol. 2020;77:1382–1389

4. Finegold K, Conmy A, Chu RC, Bosworth A, Sommers BD. Trends in the U.S. uninsured population, 2010-2020. Washington, DC: Office of the Assistant Secretary for Planning and Evaluation, U.S. Department of Health and Human Services. 2021.

5. Rae M, Copeland R, Cox, C. Tracking the rise in premium contributions and cost-sharing for families with large employer coverage. Health System Tracker. 2019.

6. Morgenstern LB, Smith MA, Sanchez BN, Brown DL, Zahuranec DB, Garcia N, Kerber KA, Skolarus LE, Meurer WJ, Burke JF, et al. Persistent ischemic stroke disparities despite declining incidence in Mexican Americans. Ann Neurol. 2013;74:778–785

7. Levine DA, Galecki AT, Okullo D, Briceño EM, Kabeto MU, Morgenstern LB, Langa KM, Giordani B, Brook R, Sanchez BN, et al. Association of blood pressure and cognition after stroke. J Stroke Cerebrovasc Dis. 2020;29:104754

8. Kleindorfer DO, Towfighi A, Chaturvedi S, Cockroft KM, Gutierrez J, Lombardi-Hill D, Kamel H, Kernan WN, Kittner SJ, Leira EC, et al. 2021 guideline for the prevention of stroke in patients with stroke and transient ischemic attack: A guideline from the American Heart Association/American Stroke Association. Stroke. 2021;52:e364–e467

9. Whelton PK, Carey RM, Aronow WS, Casey DE, Collins KJ, Dennison Himmelfarb C, DePalma SM, Gidding S, Jamerson KA, Jones DW, et al. 2017 ACC/AHA/AAPA/ABC/ACPM/AGS/APHA/ASH/ASPC/NMA/PCNA guideline for the prevention, detection, evaluation, and management of high blood pressure in adults: Executive summary: A report of the American College of Cardiology/American Heart Association Task Force on Clinical Practice Guidelines. Journal of the American College of Cardiology. 2018;71:2199–2269

10. Murao K, Bodenant M, Cordonnier C, Bombois S, Henon H, Pasquier F, Bordet R, Leys D. Does pre-existing cognitive impairment no-dementia influence the outcome of patients treated by intravenous thrombolysis for cerebral ischaemia? Journal of Neurology, Neurosurgery, and Psychiatry. 2013;84:1412–1414

11. Jorm AF. The informant questionnaire on cognitive decline in the elderly (IQCODE): A review. Int Psychogeriatr. 2004;16:275–293

12. Morales JM, Bermejo F, Romero M, Del-Ser T. Screening of dementia in community-dwelling elderly through informant report. Int J Geriatr Psychiatry. 1997;12:808–816

13. Højsgaard S, Halekoh U, Yan J. The r package geepack for generalized estimating equations. Journal of Statistical Software. 2005;15:1–11

14. Weuve J, Tchetgen Tchetgen EJ, Glymour MM, Beck TL, Aggarwal NT, Wilson RS, Evans DA, Mendes de Leon CF. Accounting for bias due to selective attrition: The example of smoking and cognitive decline. Epidemiology. 2012;23:119–23

15. van Buuren S, Groothuis-Oudshoorn K. Mice: Multivariate imputation by chained equations in r. Journal of Statistical Software. 2011;45:1–67

16. Rubin D. Multiple imputation for nonresponse in surveys. New York, NY: Wiley; 1987.

17. Sozener CB, Lisabeth LD, Shafie-Khorassani F, Kim S, Zahuranec DB, Brown DL, Skolarus LE, Burke JF, Kerber KA, Meurer WJ, et al. Trends in stroke recurrence in Mexican Americans and non-Hispanic whites. Stroke. 2020;51:2428–2434

18. Jaffe MG, Lee GA, Young JD, Sidney S, Go AS. Improved blood pressure control associated with a large-scale hypertension program. JAMA. 2013;310:699–705

19. Fletcher RD, Amdur RL, Kolodner R, McManus C, Jones R, Faselis C, Kokkinos P, Singh S, Papademetriou V. Blood pressure control among US veterans: A large multiyear analysis of blood pressure data from the Veterans Administration health data repository. Circulation. 2012;125:2462–2468

20. Shimbo D, Artinian NT, Basile JN, Krakoff LR, Margolis KL, Rakotz MK, Wozniak G. Self-measured blood pressure monitoring at home: A joint policy statement from the American Heart Association and American Medical Association. Circulation. 2020;142:e42–e63

21. Springer MV, Malani P, Solway E, Kirch M, Singer DC, Kullgren JT, Levine DA. Prevalence and frequency of self-measured blood pressure monitoring in us adults aged 50-80 years. JAMA Netw Open. 2022;5:e2231772

